# Meta-Analysis of Robustness of COVID-19 Diagnostic Kits During Early Pandemic

**DOI:** 10.1101/2021.01.16.21249937

**Authors:** Chandrakumar Shanmugam, Michael Behring, Vishwas Luthra, Sixto M Leal, Sameer Al Diffalha, Sooryanarayana Varambally, George J Netto, Upender Manne

## Abstract

**Background:** Accurate detection of severe acute respiratory syndrome corona virus 2 (SARS-CoV-2) is necessary to mitigate the coronavirus disease-19 (COVID-19) pandemic. However, the test reagents and assay platforms are varied and may not be sufficiently robust to diagnose COVID-19.

**Methods:** We reviewed 85 studies (21,530 patients), published from five regions of the world, to highlight issues involved in the diagnosis of COVID-19 in the early phase of the pandemic, following the standards outlined in the PRISMA statement. All relevant articles, published up to May 31, 2020, in PubMed, BioRiXv, MedRiXv, and Google Scholar, were included. We evaluated the qualitative (9749 patients) and quantitative (10,355 patients) performance of RT-PCR and serologic diagnostic tests for real-world samples, and assessed the concordance (5,538 patients) between methods in meta-analyses.

**Results:** The RT-PCR tests exhibited heterogeneity in the primers and reagents used. Of 1,957 positive RT-PCR COVID-19 participants, 1,585 had positive serum antibody (IgM +/- IgG) tests (sensitivity 0.81, 95%CI 0.66-.90). While 3,509 of 3581 participants RT-PCR negative for COVID-19 were found negative by serology testing (specificity 0.98, 95%CI 0.94-0.99). The chemiluminescent immunoassay exhibited the highest sensitivity, followed by ELISA and lateral flow immunoassays. Serology tests had higher sensitivity and specificity for laboratory-approval than for real-world reporting data.

**Conclusions:** The robustness of the assays/platforms is influenced by variability in sampling and reagents. Serological testing complements and may minimize false negative RT-PCR results. Lack of standardized assay protocols in the early phase of pandemic might have contributed to the spread of COVID-19.

## INTRODUCTION

In December 2019, there was a cluster of unexplained pneumonia cases in Wuhan, China, and a novel coronavirus was identified as the causative agent.^1^ The virus was named as severe acute respiratory syndrome corona virus 2 (SARS-CoV-2), and the disease as corona virus disease-19 (COVID-19).^2^ The clinical spectrum ranges from asymptomatic forms to acute respiratory failure and multi-organ dysfunction syndrome, coagulopathy, and death.^3,4^ In February 2020, the World Health Organization described the spread of these infections as a pandemic, which persists as a global crisis. Robust diagnostic tests are required to mitigate the spread of this virus and thereby to minimize the impact of COVID-19 on the health, economy, and social well-being of mankind.

The standard diagnosis of COVID-19 is based on clinical and radiologic evidence and viral genome detection by RT-PCR in respiratory samples.^5^ Gene-specific primers are used in the RT-PCR assays; structural genes include *envelope* (*E*), *nucleocapsid* (*N*), and/or *spike (S)*-genes; non-structural genes include *RNA-dependent RNA polymerase* (*RdRp*) or *open reading frame1ab* (ORF1ab) ^6,7^ Some studies used only a single-gene specific primer, and others used multiple-gene primers. Since studies published in the early phase of the pandemic reported a 3%-41% range of false-negativity by RT-PCR, a repeat RT-PCR testing was suggested.^8,9^ Furthermore, false negativity was attributed to either mutations in the regions to which the primers bind or to sampling and laboratory practices, including collection, transportation, and handling.^10^ Timing of sample collection with respect to the course of infection and the sample type also influence test results.^11^ Alternatively, the diagnosis can be made by detection of antigens (E, N, or S) and/or antibodies (IgM or IgG or both) in blood samples.^12^ However, these tests have the potential for false positives owing to cross-reactivity with other human corona viruses.^13,14^ Due to the unprecedented public health emergency, the FDA authorized, on June 1, 2020, EUA requests for more than 15 diagnostic and serologic tests. Though serology testing can detect the false positives of RT-PCR tests in clinically suspected patients, its value in COVID-19 diagnosis as a complementary assay in the mitigation of the pandemic is not well defined. However, given the complexities in COVID-19 testing, there is a need for a review of performance for tests commonly used.

In this systematic review and meta-analysis, we examine current tests for the diagnosis of COVID-19 and evaluate the sensitivity and specificity of serological tests relative to RT-PCR tests. Our objective was to identify reasons for variability in COVID-19 diagnostic tests in the early phase of the pandemic that might have contributed to the spread of COVID-19. In particular, we assessed the uniformity of primer usage in RT-PCR assays and evaluated whether primers used in gold-standard RT-PCR tests affect the validity of serological tests. Furthermore, we compared the performance of serological tests/platforms in approval contrived/laboratory *vs*. real-world data.

## METHODS

### Literature Search

This research was accomplished according to standards outlined in the Preferred Reporting Items for Systematic Reviews and Meta-Analyses (PRISMA) statement.^15^ To find relevant studies, international databases, including PubMed, MedRiXv, BioRiXv, and Google Scholar, were searched for articles published until May 31, 2020. The following search terms were used (selected using English MeSH keywords and Emtree terms): [SARS-CoV-2 AND diagnosis] OR [2019-nCoV AND diagnosis]” OR [“COVID-19 AND diagnosis] and [SARS-CoV-2 AND RT-PCR] OR, [2019-nCoV AND RT-PCR]” OR [“COVID-19 AND RT-PCR] and [SARS-CoV-2 AND serology] OR [2019-nCoV AND serology]” OR [“COVID-19 AND serology]. Additional searches were performed for references listed in the included studies.

### Eligibility Criteria

Relevant articles that reported diagnostic information for infected patients were included in the analysis. Pre-print articles with non-peer review were considered for inclusion. Articles were excluded if appropriate information was not reported or if they were in the Chinese language. Population sample sizes of <5 participants were not included; reviews and editorials were not considered. For meta-analysis and approval *vs*. real-world performance, studies that reported percent sensitivity/specificity without including patient numbers were also excluded.

### Data Extraction and Report Quality Evaluation

Two authors (CS and VL) screened and evaluated the literature independently. Discrepancies were resolved by consensus after evaluation by a third author (MB). The following were extracted for review and meta-analysis: journal name, authors, period of publication (end of May, 2020), location of study, total number of patients, tissue of origin for samples tested, whether samples were from upper or lower respiratory tract (or both), primers for RT-PCR, platforms for serology tests, and antibodies tested for serology. Counts of true positives, false negatives, true negatives, and false positives were used in the meta-analysis.

An author (MB) extracted and analyzed the approved testing kit performance data from the following sources: FDA EUA Authorized Serology Test Performance,^16^ the Foundation for Innovative New Diagnostics (FIND) SARS-CoV-2 diagnostic pipeline,^17^ and package inserts provided on company websites for each product. Real-world sample testing data from kits in meta-analyses were compared against the performance of the same kits, or platforms, reported in approval documentation. Variables abstracted were study authors/test developer, name of test, test platform, and true positives, false negatives, true negatives, and false positives for each antibody or antibody combination measured (IgM, IgG, IgA, combined, and Pan-Ig). Risk of bias within individual studies of meta-analysis was assessed using the QUADAS 2 tool for assessment of diagnostic studies.^18^

### Statistical Analysis

Statistical analyses were performed with R version 6.3.2 (2019-12-12).^19^ The package “meta” was used for meta-analyses.^20^ Random effects models were used to measure sensitivity and specificity of outcomes across studies. Subgroup analysis was performed to evaluate the effect of assay, RT-PCR primer type, and setting (laboratory *vs*. real-world) upon serum test performance. Heterogeneity across studies and subgroups was evaluated using Cochrane’s *Q* statistic, and residual heterogeneity was quantified as a percentage with the *I*^*2*^ statistic. An *I*^*2*^ measure of 0% shows no observed heterogeneity, with increasing values from 0%-100% indicating higher levels of heterogeneity. ^21^ An assumption of homogeneity was rejected for p-values < 0.1. Evaluation of publication bias was not possible in approval data.

## RESULTS

### Search Results and Population Characteristics

Our search generated 112 publications with potential relevance to the performance of COVID-19 diagnostic tests. After excluding duplicate publications, manuscripts that did not report numbers of patients used for sensitivity/specificity calculations and studies with a sample size of <5 patients, 85 studies were selected for qualitative synthesis of RT-PCR primer usage. From this set, a sub-set of 30 publications were selected for the quantitative meta-analysis of serologic *vs*. RT-PCR diagnostic testing for COVID-19 (**Table S1**). Ancillary analysis compared the performance of these 30 real-world studies to that reported in laboratory approval data from 47 diagnostic serum-based tests. In all, our qualitative synthesis of RT-PCR studies included 85 studies and 21,530 patients. From this synthesis, a group of 30 studies with 10,355 patients from 5 regions of the world were selected for meta-analysis and comparison to performance from laboratory approval data (**Fig S1**).

### Uniformity of Primer Usage in RT-PCR Diagnostic Tests

We reviewed use of single primer of structural genes as compared to use of both structural and non-structural gene primers in 56 population-based studies with 9,872 participants. Overall, high proportions of studies employed both structural and non-structural gene primers in RT-PCR testing (58% in studies and 56% of total participants). Additionally, 29 studies (11,658 patients) did not report RT-PCR primer data. Single markers were most frequently tested in China and North American studies (**Table 1**). In general, the most tested samples were from the upper respiratory tract, regardless of primer status. Sample source and location in the respiratory tract were not reported for 8-20% of patients, and this was more common for studies using single gene primer.

**Table 1.** Characteristics of the studies included in qualitative analysis.

|  | Total Studies | Total pop. | Structural gene primers |  | Structural and non-Structural gene primers |  | Non-Structural gene primers |  | Not reported |  |
| --- | --- | --- | --- | --- | --- | --- | --- | --- | --- | --- |
|  |  |  | N studies | N pop. | N studies | N pop. | N studies | N pop. | N studies | N pop. |
| **Total** | **85** | **21530** | **22** | **4265** | **31** | **5484** | **3** | **123** | **29** | **11658** |
| **Location** |  |  |  |  |  |  |  |  |  |  |
| Asia (excl. China) | 6 | 378 | 2 | 53 | 4 | 325 |  |  |  |  |
| China | 28 | 12187 | 8 | 1802 | 17 | 3047 | 3 | 123 | 24 | 7215 |
| Europe | 12 | 5757 | 4 | 528 | 8 | 993 |  |  | 4 | 4236 |
| North America | 10 | 3001 | 8 | 1882 | 2 | 1119 |  |  |  |  |
| Global |  | 207 |  |  |  |  |  |  | 1 | 207 |
| **Primers** |  |  |  |  |  |  |  |  |  |  |
| N -single | 11 | 2016 | 11 | 2016 |  |  |  |  |  |  |
| E -single | 4 | 759 | 4 | 759 |  |  |  |  |  |  |
| S -single | 1 | 412 | 1 | 412 |  |  |  |  |  |  |
| N, E | 2 | 226 | 2 | 226 |  |  |  |  |  |  |
| S, N | 4 | 852 | 4 | 852 |  |  |  |  |  |  |
| ORF1Ab, single | 2 | 59 |  |  |  |  | 2 | 59 |  |  |
| RdRp, single | 1 | 64 |  |  |  |  | 1 | 64 |  |  |
| E+ORF1Ab | 2 | 1119 |  |  | 2 | 1119 |  |  |  |  |
| E + RdRp | 2 | 259 |  |  | 2 | 259 |  |  |  |  |
| M, E | 1 | 48 |  |  | 1 | 48 |  |  |  |  |
| N+ORF1Ab | 14 | 2703 |  |  | 14 | 2703 |  |  |  |  |
| N + E + RdRp | 4 | 333 |  |  | 4 | 333 |  |  |  |  |
| S, N, E, RdRp, ORF1ab | 1 | 13 |  |  | 1 | 13 |  |  |  |  |
| N, E, ORF1ab | 1 | 33 |  |  | 1 | 33 |  |  |  |  |
| N, RNase P | 1 | 190 |  |  | 1 | 190 |  |  |  |  |
| S, N, RdRp, ORF1ab, E, M | 1 | 52 |  |  | 1 | 52 |  |  |  |  |
| N, S, RdRp | 1 | 273 |  |  | 1 | 273 |  |  |  |  |
| N, E, S, RdRp | 2 | 349 |  |  | 2 | 349 |  |  |  |  |
| S, ORF1Ab | 1 | 112 |  |  | 1 | 112 |  |  |  |  |
| **Sample Source** |  |  |  |  |  |  |  |  |  |  |
| Upper Respiratory | 23 | 6748 | 3 | 575 | 9 | 2633 | 1 | 64 | 10 | 3476 |
| Upper & Lower Respiratory | 1 | 52 |  |  | 1 | 52 |  |  |  |  |
| Upper Respiratory + Other* | 9 | 751 | 3 | 368 | 2 | 44 | 1 | 38 | 3 | 301 |
| Lower Respiratory + Other* | 1 | 273 |  |  | 1 | 273 |  |  |  |  |
| Upper Respiratory + Serum | 20 | 6407 | 7 | 1473 | 9 | 1432 |  |  | 4 | 3502 |
| Upper Respiratory + Serum + Other* | 4 | 941 | 2 | 840 | 1 | 80 | 1 | 21 |  |  |
| Upper & Lower Respiratory + Other* | 4 | 678 | 1 | 280 | 3 | 398 |  |  |  |  |
| Upper & Lower Respiratory + Serum + Other* | 2 | 518 |  |  | 1 | 132 |  |  | 1 | 386 |
| Serum | 18 | 2376 | 6 | 729 | 4 | 440 |  |  | 8 | 1207 |
| Other* | 1 | 199 |  |  |  |  |  |  | 1 | 199 |
| Not reported | 2 | 2587 |  |  |  |  |  |  | 2 | 2587 |

\* Other = bronchioalveolar lavage, feces, urine, neonatal, amniotic fluid, and breast milk. N pop. = patient population

### Meta-Analysis: RT-PCR *vs*. Serum Antibody Testing

In general, patient sera were tested for IgM and IgG antibodies. China was the region with the highest frequency of antibody testing, and lateral flow immunoassay (LFIA) and chemiluminescent immunoassay (CLIA) testing platforms were most often utilized. Of the 45 studies included in the qualitative synthesis, 30 manuscripts reported both serum antibody testing and RT-PCR testing for the same patients. Key characteristics of this population include: China as the regional location for research; lack of reporting of RT-PCR primer information for ∼33% of all studies; most studies used IgM and IgG serum-based antibody tests; and LFIA, CLIA, and enzyme-linked immunosorbent assay (ELISA) platforms were common across studies (**Table 2**).

**Table 2.**
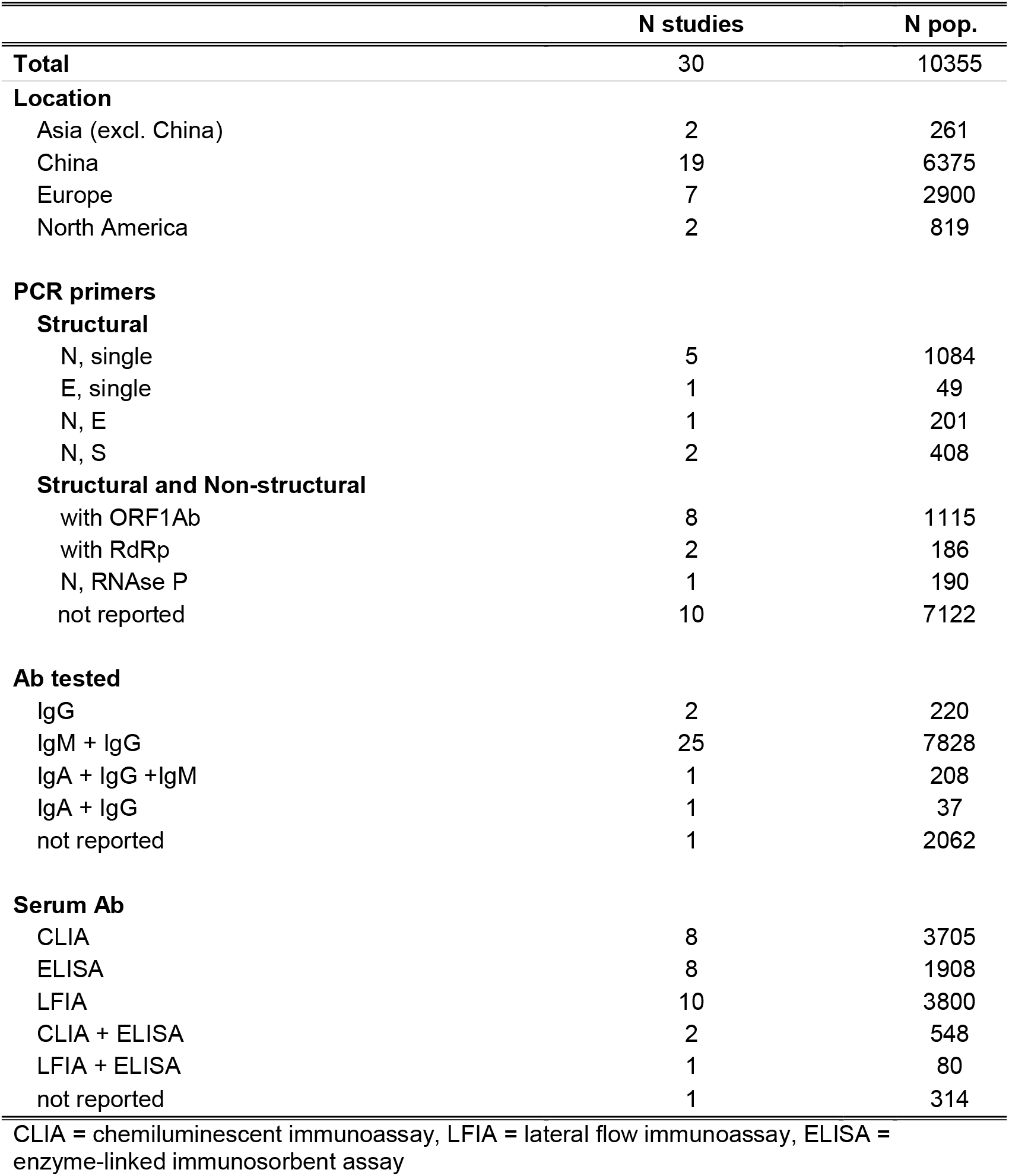
Characteristics of studies included in quantitative meta-analysis.

We used the IgM+/-IgG serum antibody test since it was most commonly utilized across studies. Of 1,957 participants (sensitivity 0.81, 95%CI 0.66-0.90) with a positive RT-PCR COVID-19 result, 1,585 were also detected as positive with serum antibody tests. Of 3,581 true negatives in RT-PCR, 3,509 negatives were also found by serum antibody testing (specificity 0.98, 95%CI 0.94-0.99). For both models, heterogeneity between studies was significant (p<0.01 for both, *I*^*2*^=97% and *I*^*2*^ =98% for sensitivity and specificity, respectively).

Sub-analyses of differences based on the testing platform found that sensitivity between groups differed (p <0.0001), with CLIA tests performing best (0.99, 95%CI 0.97-0.99); ELISA as next-best (0.89, 95%CI 0.82-0.93); and LFIA as having the poorest sensitivity (0.67, 95%CI 0.50-0.81). LFIA test sensitivity also showed heterogeneity between studies (p<0.01, *I*^*2*^ 95%). For IgM/IgG tests, specificity did not differ significantly by platform (p= 0.06). However, a performance trend followed sensitivity, with LFIA underperforming (**Figure 1**).

**Figure 1.**
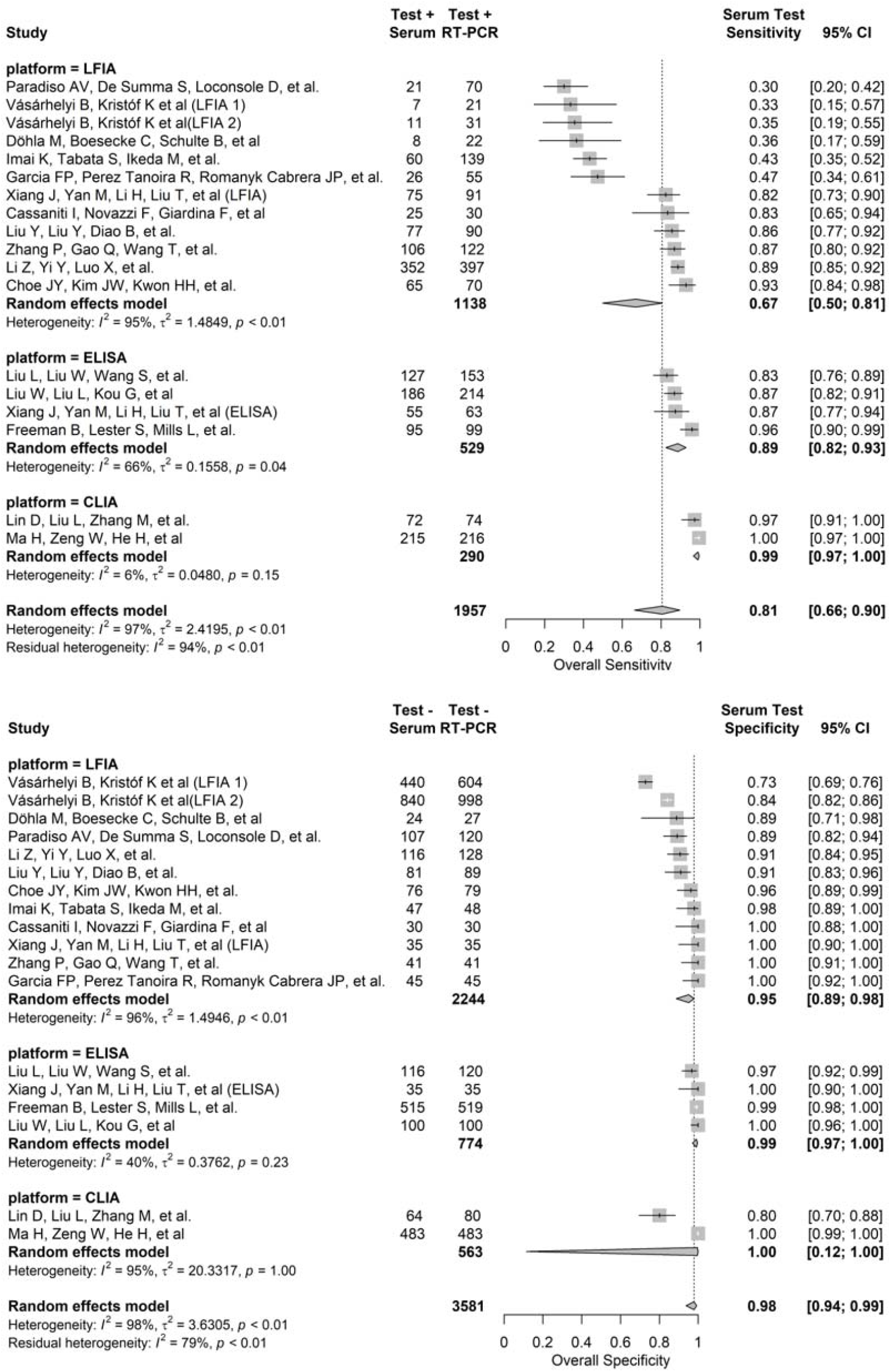
Comparison of Performance (Sensitivity and Specificity) of Serology Tests (IgM/IgG) Based on Assay Platforms.

### Serum Antibody Testing Performance: Approval Data vs. Real-World Data

For manufacturer-based, laboratory approval data, IgM+/-IgG testing detected COVID-19 positivity for 1,045 of 1,068 RT-PCR-determined “true” positive patients (sensitivity 0.98, 95%CI 0.92-1.0). In the same group, serum testing correctly identified 1,928 of 1,967 (specificity 0.98, 95%CI 0.95-.099) true negatives by RT-PCR. For both models (sensitivity and specificity), there was evidence of heterogeneity (p <0.01 for both and *I*^*2*^=93% and *I*^*2*^=94% for sensitivity and specificity, respectively).

We evaluated IgM+/-IgG serum test performance in subgroup analyses comparing laboratory approval performance data to real-world performance in study data. In manufacturer data presented for approval, serum antibody testing detected 1,047 of 1,068 “true positive” cases of COVID-19 (sensitivity 0.98, 95%CI 0.92-1.0). Real-world use of serum IgM+/-IgG testing was evident for 2,450 of 3,025 participants diagnosed with COVID-19 by RT-PCR (sensitivity 0.81, 95% CI 0.66-0.90). For both groups, there was heterogeneity between studies (p <0.01 for both, *I*^*2*^=93% and *I*^*2*^ =97% for approval and real-world specificity, respectively) (**Figure 2**). In addition, the overall sensitivity between approval and real-world testing groups differed significantly (*Q*=8.37, p=0.004). An analysis of specificity by the same subgroups found no significant difference between laboratory approval and real-world data. Laboratory data identified 1,928 of 1,967 participants with true COVID-19 negative status (specificity 0.98, 95% CI 0.95-0.99). Real-world data found 5,437 of 5,548 true negatives (specificity 0.98, 95% CI, 0.96-0.99) (analysis not shown).

**Figure 2.**
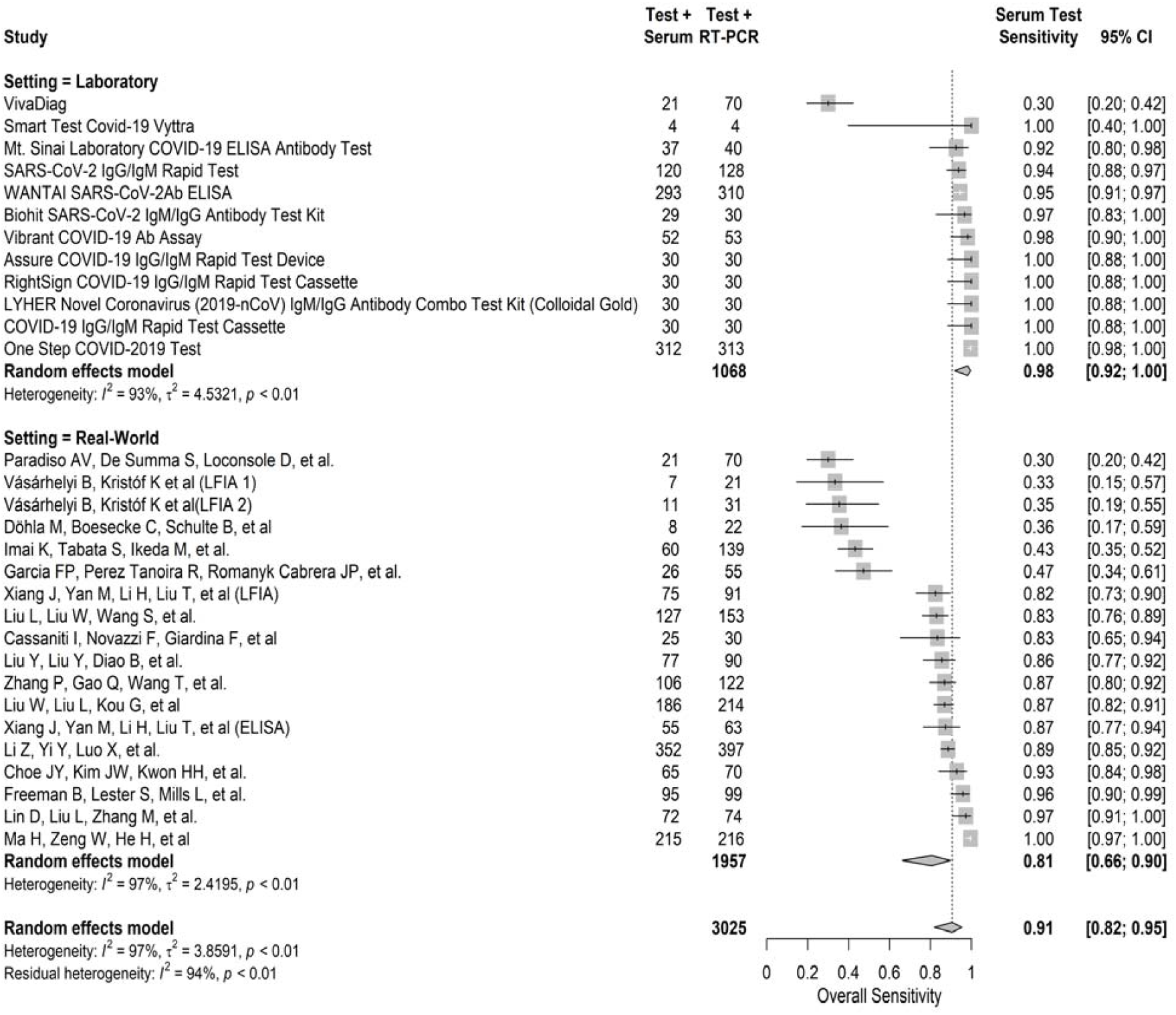
Comparison of Sensitivity of Laboratory setting versus Real World setting of RT-PCR and Serology (IgM/IgG) kits.

Since, in IgM+/-IgG tests, there were differences in sensitivity between platforms, we evaluated the effect of approval-based data *vs*. real-world data by the type of platform. In an analysis stratified for ELISA, CLIA, and LFIA, there was no significant difference in specificity between approval and real-world data (data not shown). However, for ELISA tests, real-world capacity to detect true positives was lower than in laboratory-based analyses. In real-world studies, the sensitivity of ELISA was 0.89 (95% CI, 0.82-0.93), different from laboratory sensitivity for the same platform (0.94, CI95% 0.91-0.96, *Q* =4.74, p=0.03). The LFIA platform also showed a trend of lower real-world sensitivity (0.67, 95% CI, 0.50-0.81) compared to laboratory approval sensitivity (0.99, CI95% 0.90-0.99, *Q* =8.56, p 0.003). Laboratory/real-world groups for CLIA platforms were too small to be tested reliably (1 and 2 groups, respectively).

### Serum Antibody Testing Performance: Effect of Primer Choice on Test Validity

To evaluate the reliability of RT-PCR as a gold standard for serum-based test performance, we tested the consequences of using structural and non-structural primers in RT-PCR reference testing of serum. Analyses were divided into three subgroups based on antibody targets: IgM, IgG, and IgG+/-IgM combined. In IgM and combined IgG+/-IgM testing, the primer choice had no significant influence on sensitivity or specificity. However, for IgG antibody tests, use of both a structural and a non-structural gene-specific primers in RT-PCR resulted in reduced sensitivity for serum testing (**Figure 3**, *Q*=6.17, p=0.013). Furthermore, although not statistically significant, the sensitivity of both IgM and IgG+/-IgM combined data sets was lower when using a referent RT-PCR test with both primer types.

**Figure 3.**
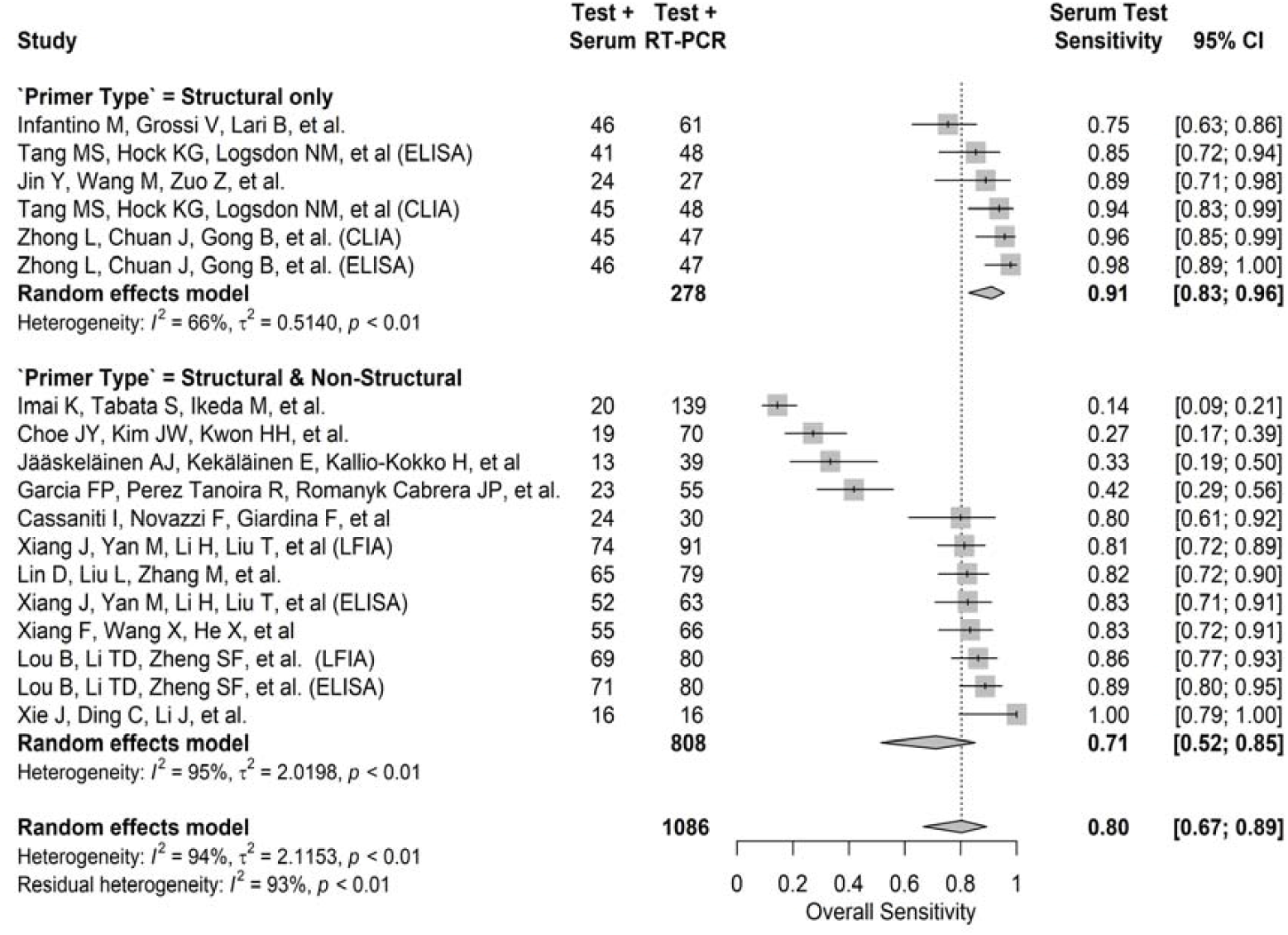
The Effect of Primer Choice in RT-PCR Referent on Sensitivity of Tests based on Serum IgG.

## DISCUSSION

Because of the highly infectious nature of COVID-19, a prompt, accurate, and early diagnosis is necessary to deal with the ongoing pandemic, for such diagnoses can help reduce the spread of infection and its associated risk for mortality. Currently, the COVID-19 diagnosis is generally based on RT-PCR assays.^8^ Alternative methods such as antigen- and antibody-based serology tests, although available, have uncertain value. The current systematic review and meta-analysis addresses the challenges encountered in the diagnosis of COVID-19 by various methods. It also analyzes differences between the FDA-approved EUA data and real-world data. There is worldwide non-uniformity in the performance of RT-PCR, including the number and types of primers and reagents used for COVID19 diagnosis, which raise questions about its generalized applicability. Similarly, the studies based on serological tests showed diagnostic inaccuracies owing to individual differences in mounting an immune response as well as dependency on the time duration after the onset of symptoms. Overall, the sensitivity between RT-PCR and serology tests was 0.81 (95% CI, 0.66-0.90), and specificity was 0.98 (95% CI, 0.94-0.99). Among the various platforms for serodiagnosis, the highest sensitivity was exhibited by ELISA, followed by CLIA and LFIA. Furthermore, use of primers (structural, non-structural, or both) had a variable effect on sensitivity based on antibody targets. Sensitivity was significantly higher for IgG serology tests using structural-primer-only RT-PCR tests as a referent. Serology tests had higher sensitivity for approval-based data than for real-world reporting. This difference was significant for ELISA-based platforms, and a non-significant trend towards inflated approval-based sensitivity was evident for both CLIA and LFIA platforms. These observations highlight the inconsistencies/challenges in the COVID-19 diagnosis by RT-PCR, which is the current gold standard, as well as in serologic testing.

For RT-PCR assays, the targets in SARS-CoV-2 include structural genes like *E, N* and *S*, and nonstructural genes, including that for *RdRp* or *ORF1ab*.^22^ In the early phase of the pandemic, some studies used a two-step diagnosis that included an initial screening phase using structural genes followed by a confirmatory phase using nonstructural genes. ^6,7,23^ The test is considered positive when both structural and non-structural markers are positive.^24,25^ However, currently both types of primers are used simultaneously to diagnose COVID-19. The viral load or copy number of the viral genome is expressed as a Ct-value, which when <37 is indicative of a positive test, and a value of ≥ 40 is considered negative. A Ct value between >37 and < 40 requires repetition of RT-PCR analysis to confirm the diagnosis.^24^ However, the Ct value range varies widely according to assays and laboratory practices. A COVID-19-RdRp/Hel assay has a higher sensitivity than a conventional RdRp-P2 assay irrespective of the type of sample.^26^ Overall, higher proportions of studies (58%) employed both structural and non-structural gene primers in RT-PCR testing. Single markers were used in some Chinese and North American studies. These findings are indicative of non-uniformity in the RT-PCR methodology. We note that half of the positive, symptomatic patients became negative by the second week, when they became asymptomatic. In contrast, the asymptomatic, positive patients became negative two days after hospital admission, indicating the importance of a temporal factor in COVID-19 diagnosis by RT-PCR.^27,28^

Published in the early phase of the pandemic, 11 of 85 studies had clinically suspected COVID-19 patients. In these studies, the average test positivity by RT-PCR, regardless of the sample source, was 44% (Supplementary Table 1), and test sensitivity was influenced by sample source (upper *vs*. lower respiratory *vs*. other samples), issues related to testing performance, and delay after onset of symptoms.^29^ In the early phase of the COVID-19 pandemic, for studies evaluating suspected COVID-19 cases, the total positive RT-PCR for throat swabs was in the range of 30–60% at initial presentation.^8,30^ One study reported a yield of 72-93% positive cases for lower respiratory samples (bronchioalveolar lavage and sputum) as compared to 32-63% positivity for upper respiratory samples (oral and nasopharyngeal swabs) and 29% for stool samples.^29^ Hence, a negative COVID-19 test based only on an upper respiratory sample at a single time point is questionable. For most studies, the testing sample was from the upper respiratory tract, regardless of primer type used. However, the sample source was not reported for 8-20% of patients, which was more common for studies using only structural gene primers. For stool samples testing positive for COVID-19, 66.7% also tested positive on pharyngeal swabs. Of the stool samples, 64.3% remained positive after pharyngeal clearance of the virus.^31^ In contrast, none of the patients showed a positive test on upper respiratory samples after the anal swabs tested negative.^31^ These findings raise concerns about whether patients with negative respiratory swabs are truly virus-free, and sampling of additional body sites is needed. As determined by various studies, the performance of the RT-PCR depends on usage of comparable protocols, including primers and reagents.^32^ Additionally, it is unknown whether the currently used RT-PCR primers detect all SARS-Cov-2 strains.

The specific immune response to SARS-CoV-2 can be measured by serological testing. Several rapid serological tests, including point-of-care tests, are being developed. Even though some of these tests have been approved by the FDA through EUA, their accuracy needs to be validated.^33^ A minimum of 1–2 weeks after the onset of infection is needed for seroconversion. Hence, antibody testing is of no value in the early phase of infection. Additionally, its value is limited by its cross-reactivity with other coronaviruses.^34,35^ The initial RT-PCR positivity during the early stages (<15 days) of SARS-CoV-2 infection declines to 66.7% in the later phase (15-39 days), during this period, the antibody test can supplement RT-PCR in the diagnosis of COVID-19.^34,35^ Additionally, serology testing becomes valuable for clinically suspected and RT-PCR negative (false-negative) individuals.

This research has limitations. Due to the dynamic reporting of COVID-19 testing research and inconsistencies in reporting of predictive variables across studies, bias in sampling may have some effect on our results. Patient flow analysis suggests that lack of consistent RT-PCR reference standard given to patients in the same study, as well as the unclear reporting of patient selection methods could contribute to bias in these results (**Fig. S2**). In addition, the observed heterogeneity between studies in the meta-analysis suggests that we must consider the possibility that the differences in results may be due to chance. Lastly, it is questionable to compare two separate testing methods of RT-PCR and seroprevalence in sensitivity/specificity analysis. In particular, given the relationship between time since diagnosis and accuracy of serology testing, a contributor to the observed differences in performance is time.

The effective containment of COVID-19 involves accurate diagnoses and isolation of SARS-CoV-2-infected persons. Robustness of the assays/platforms is determined by variability of the samples, primers, and reagents used. Serological tests alone are of value only during the latter times of infection; however, they complement RT-PCR when used in conjunction and minimize false negative RT-PCR results. Additionally, some of the approved serological assays/platforms, particularly those developed using contrived/laboratory data, perform poorly when applied to real-world samples. We are currently in a new phase of the pandemic, and there is a need for a reliable/robust diagnostic test to mitigate the spread of COVID-19.

Our analyses of studies published in the early-phase of the pandemic have highlighted issues related to COVID-19 diagnosis that need to be addressed as follows: 1) The high mutational rate exhibited by the SARS-CoV-2 virus may lead to the generation of new strains. Therefore, like for influenza virus, the existing diagnostic kits need to be modified constantly to optimize the detection of new strains; 2) Though RT-PCR diagnosis of COVID-19 is the gold standard, its combination with a serologic test may increase the accuracy of SARS-CoV-2 detection; 3) Approval agencies must account for the type of data (contrived versus real world) presented by diagnostic kit developer; 4) Although agencies employed EUA processes for the approval of diagnostic kits, there is a need to monitor their performance and assess their robustness in real-world samples, to permit continued use of these kits; and 5) Standardized assay protocols need to be developed and continually updated to mitigate the COVID-19 pandemic.

## Data Availability

All data is publicly available as detailed in supplementary table 1

## ACKNOWLEDGEMENTS

This study was supported, in part, by grant 5U54CA118948 and institutional/departmental impact funds (UM), and by CA047888, the UAB Cancer Prevention and Control Training Program (MB). We thank Dr. Donald Hill for his editorial assistance.

