## Supplemental Information for "Meta-Analysis of Robustness of COVID-19 Diagnostic Kits During Early Pandemic"

**Table S1. Description of 55 Studies Included for Qualitative Synthesis (gray) and 30 Studies Included in the Meta-Analysis (tan)**

| **Journal** | **Authors** | **Region** | **N (patients)** | **Percent COVID-19 positivity** | **Sample origin** | **RT-PCR primers** | **RT-PCR primer type** | **RT-PCR/Serology platform** | **Serology targeted antibody** | **Study included in meta-analysis** |
| --- | --- | --- | --- | --- | --- | --- | --- | --- | --- | --- |
| *J Clin Virol* | Rahman H, Carter I, Basile K, et al. ^1^ | Asia | 52 |  | UR+LR | S, N, E, RdRp, ORF1ab, M | Both | RT-PCR |  | No |
| *Exp Neurobiol* | Won J, Lee S, Park M, et al. ^2^ | Asia | 12 |  | UR | N, E, S, RdRp | Both | RT-PCR |  | No |
| *Jpn J Infect Dis.* | Okamaoto K, Shirato K, Nao N, et al. ^3^ | Asia | 25 |  | UR | N, E | Structural | RT-PCR |  | No |
| *J Med Virol* | Choe JY, Kim JW, Kwon HH, et al. ^4^ | Asia | 149 | 0.47 | Serum | E, RdRp | Both | RT-PCR /CLIA | IgM + IgG | Yes |
| *Lancet Infect Dis* | Yong SEF, Anderson DE, Wei WE, et al. ^5^ | Asia | 28 |  | UR + Serum | N, single | Structural | RT-PCR /ELISA | IgG | No |
| *J Clin Virol* | Imai K, Tabata S, Ikeda M, et al. ^6^ | Asia | 112 |  | UR + Serum | S, ORF1Ab | Both | RT-PCR /LFIA | IgM + IgG | Yes |
| *Emerg Microbes Infect* | Xu Y, Xiao M, Liu X, et al. ^7^ | China | 6 |  | Serum |  |  | ELISA + LFIA | IgM | No |
| *Radiology* | Ai T, Yang Z, Hou H, Zhan C, et al. ^8^ | China | 1014 | 0.59 | UR |  |  | RT-PCR |  | No |
| *NEJM* | Cao B, et al.^9^ | China | 199 |  | Other |  |  | RT-PCR |  | No |
| *Radiology* | Bai HX, Hsieh B, Xiong Z, et al.^10^ | China | 256 |  | UR |  |  | RT-PCR |  | No |
| *Lancet* | Chen H, Guo J, Wang C, et al. ^11^ | China | 9 |  | UR + Other |  |  | RT-PCR |  | No |
| *AJR Am J Roentgenol* | Liu D, Li L, Wu X, et al. ^12^ | China | 15 |  | UR |  |  | RT-PCR |  | No |
| *Eur J Radiol* | Long C, Xu H, Shen Q, et al. ^13^ | China | 87 |  | UR |  |  | RT-PCR |  | No |
| *Pediatr Pulmonol* | Xia W, Shao J, Guo Y, Peng X, Li Z, Hu D. ^14^ | China | 20 |  | UR |  |  | RT-PCR |  | No |
| *Am J Obstetr Gynecol* | Yan J, Guo J, Fan C, et al. ^15^ | China | 116 | 0.56 | Other |  |  | RT-PCR |  | No |
| *J Hosp Infect* | Ye G, Li Y, Lu M, et al. ^16^ | China | 91 | 0.52 | UR |  |  | RT-PCR |  | No |
| *J Med Virol* | Zhang J, Wang S, Xue Y. ^17^ | China | 14 |  | UR + Other |  |  | RT-PCR |  | No |
| *Respir Res* | Zhang G, Zhang J, Wang B, Zhu X, Wang Q, Qiu S. ^18^ | China | 95 |  | UR |  |  | RT-PCR |  | No |
| *Lancet* | Zhou F, Yu T, Du R, et al. ^19^ | China | 191 |  | UR |  |  | RT-PCR |  | No |
| *J Clin Microbiol* | Liu W, Liu L, Kou G, et al.^20^ | China | 314 |  | UR + Serum |  |  | RT-PCR | IgM + IgG | Yes |
| *J Med Virol* | Li, Y et al. ^21^ | China | 610 | 0.40 | UR | N, ORF1Ab | Both | RT-PCR |  | No |
| *medRxiv* | Diao B, Wen K, Chen J, et al.^22^ | China | 239 |  | UR + Serum | N, ORF1Ab | Both | RT-PCR |  | No |
| *J Clin Microbiol* | Chan JF, Yip CC, To KK, et al. ^23^ | China | 273 |  | UR + Other | N, S, RdRp | Both | RT-PCR |  | No |
| *Nature Microbiol* | Kong WH, Li Y, Peng MW, et al. ^24^ | China | 640 |  | UR | N, ORF1Ab | Both | RT-PCR |  | No |
| *Front Med* | Liu W, Wang J, Li W, Zhou Z, Liu S, Rong Z. ^25^ | China | 38 | 0.53 | UR + Other | N, ORF1Ab | Both | RT-PCR |  | No |
| *Int J Biol Sci* | Lo IL, Lio CF, Cheong HH, et al. ^26^ | China | 10 |  | UR + LR + Other | N, ORF1Ab | Both | RT-PCR |  | No |
| *Travel Med Infect Dis* | Wu J, Liu J, Li S, Peng Z, et al. ^27^ | China | 132 |  | UR + LR + Serum + Other | N, E, RdRp | Both | RT-PCR |  | No |
| *Int J Infect Dis* | Xu T, Chen C, Zhu Z, et al. ^28^ | China | 51 |  | UR + LR + Other | N, ORF1Ab | Both | RT-PCR |  | No |
| *J Med Virol* | Yuan Y, Wang N, et al. ^29^ | China | 6 |  | UR + Other | N, E, RdRp | Both | RT-PCR |  | No |
| *AJR Am J Roentgenol* | Cheng Z, Lu Y, Cao Q, et al. ^30^ | China | 33 | 0.33 | UR | N, E, ORF1ab | Both | RT-PCR |  | No |
| *Arch Pathol Lab Med* | Schwartz, DA ^31^ | China | 38 |  | UR + Other | ORF1Ab, single | Non-structural | RT-PCR |  | No |
| *Radiology* | Wong HYF, Lam HYS, Fong AH, et al. ^32^ | China | 64 |  | UR | RdRp, single | Non-structural | RT-PCR |  | No |
| *Chin Med J* | Ling Y, Xu SB, Lin YX, et al. ^33^ | China | 292 |  | UR + Other | E, single | Structural | RT-PCR |  | No |
| *Clin Infect Dis* | Zhao R, Li M, Song H, et al. ^34^ | China | 412 |  | UR | S, single | Structural | RT-PCR |  | No |
| *medRxiv* | Ma H, Zeng W, He H, et al.^35^ | China | 699 |  | UR + Serum |  |  | RT-PCR /CLIA | IgM + IgG | Yes |
| *medRxiv* | Cai X, Chen J, Hu J, et al.^36^ | China | 443 |  | Serum |  |  | RT-PCR /CLIA | IgM + IgG | Yes |
| *medRxiv* | Qian C, Zhou M, Cheng F, et al. ^37^ | China | 2062 |  |  |  |  | RT-PCR /CLIA | IgM + IgG | Yes |
| *J Infect Dis* | Zhang G, Nie S, Zhang Z, Zhang Z. ^38^ | China | 112 |  | UR + Serum | N, ORF1Ab | Both | RT-PCR /CLIA | IgM + IgG | No |
| *medRxiv* | Lin D, Liu L, Zhang M, et al.^39^ | China | 159 |  | UR + Serum | N, ORF1Ab | Both | RT-PCR /CLIA | IgM + IgG | Yes |
| *J Med Virol* | Xie J, Ding C, Li J, et al. ^40^ | China | 56 |  | UR +Serum | N, ORF1Ab | Both | RT-PCR /CLIA | IgM + IgG | Yes |
| *Nature Med* | Long QX, Liu BZ, Deng HJ, et al. ^41^ | China | 285 |  | UR+ Serum | S, N | Structural | RT-PCR /CLIA | IgM + IgG | No |
| *Int J Infect Dis* | Jin Y, Wang M, Zuo Z, et al. ^42^ | China | 76 | 0.57 | Serum | N, single | Structural | RT-PCR /CLIA | IgM + IgG | Yes |
| *Emerg Microbes Infect* | Zhang W, Du RH, Li B, et al. ^43^ | China | 278 |  | UR + Other |  |  | RT-PCR /ELISA | IgM + IgG | No |
| *Clin Infect Dis* | Zhao J, Yuan Q, Wang H, et al.^44^ | China | 386 |  | UR + LR + Serum |  |  | RT-PCR /ELISA | IgM + IgG | Yes |
| *Euro Surveill* | Perera RA, Mok CK, Tsang OT, et al. ^45^ | China | 51 |  | Serum |  |  | RT-PCR /ELISA | IgM + IgG | Yes |
| *Clin Infect Dis* | Xiang F, Wang X, He X, et al. ^46^ | China | 216 |  | UR + Serum | N, ORF1Ab | Both | RT-PCR /ELISA | IgM + IgG | Yes |
| *medRxiv* | Xiang J, Yan M, Li H, Liu T, et al. ^47^ | China | 154 |  | Serum | N, ORF1Ab | Both | RT-PCR /ELISA | IgM + IgG | Yes |
| *medRxiv* | Liu L, Liu W, Wang S, et al.^48^ | China | 238 |  | UR + Serum | N, ORF1Ab | Both | RT-PCR /ELISA | IgM + IgG | Yes |
| *Clin Infect Dis* | Guo L, Ren L, Yang S, et al. ^49^ | China | 208 | 0.39 | Serum | N, single | Structural | RT-PCR /ELISA | IgM + IgA + IgG | Yes |
| *Sci China Life Sci* | Zhong L, Chuan J, Gong B, et al.^50^ | China | 347 |  | UR NP/OP + Serum + Other | N, S | Structural | RT-PCR /ELISA + CLIA | IgM + IgG | Yes |
| *Eur Respir J* | Lou B, Li TD, Zheng SF, et al. ^51^ | China | 80 |  | UR + LR + Serum + Other | N, ORF1Ab | Both | RT-PCR /ELISA + LFIA + CLIA | IgM + IgG | Yes |
| *J Med Virol* | Du Z, Zhu F, Guo F, Yang B, Wang T. ^52^ | China | 60 |  | Serum |  |  | RT-PCR /LFIA | IgM + IgG | No |
| *J Infect* | Pan Y, Li X, Yang G, et al. ^53^ | China | 105 |  | Serum |  |  | RT-PCR /LFIA | IgM + IgG | No |
| *J Med Virol* | Li Z, Yi Y, Luo X, et al.^54^ | China | 525 |  |  |  |  | RT-PCR /LFIA | IgM + IgG | Yes |
| *medRxiv* | Liu Y, Liu Y, Diao B, et al.^55^ | China | 179 |  | UR + Serum |  |  | RT-PCR /LFIA | IgM + IgG | Yes |
| *Emerg Microbes Infect* | Yongchen Z, Shen H, Wang X, et al. ^56^ | China | 21 |  | UR + Serum + Other | ORF1Ab, single | Non-structural | RT-PCR /LFIA | IgM + IgG | No |
| *Anal Chem* | Chen Z, Zhang Z, Zhai X, et al.^57^ | China | 19 |  | UR + Serum | N, single | Structural | RT-PCR /LFIA | IgG | Yes |
| *medRxiv* | Zhang P, Gao Q, Wang T, et al.^58^ | China | 163 |  | UR + Serum | N, single | Structural | RT-PCR /LFIA | IgM + IgG | Yes |
| *JAMA* | Grasselli G, Zangrillo A, Zanella A, et al. ^59^ | Europe | 1591 |  | UR |  |  | RT-PCR |  | No |
| *Radiology* | Caruso D, Zerunian M, Polici M, et al. ^60^ | Europe | 158 | 0.39 | UR | N, E, RdRp | Both | RT-PCR |  | No |
| *Travel Med Infect Dis* | Lagier JC, Colson P, Tissot Dupont H, et al. ^61^ | Europe | 337 |  | UR +LR+ Other | N, E, S, RdRp | Both | RT-PCR |  | No |
| *J Clin Virol* | van Kasteren PB, van der Veer B, van den Brink S, et al. ^62^ | Europe | 13 |  | UR | S, N, E, RdRp, ORF1ab | Both | RT-PCR |  | No |
| *Int J Mol Sci* | Toptan T, Hoehl S, Westhaus S, et al. ^63^ | Europe | 48 |  | UR | M, E | Both | RT-PCR |  | No |
| *Trop Med Infect Dis* | Amrane S, Tissot-Dupont H, Doudier, et al. ^64^ | Europe | 280 |  | UR + LR + Other | E, single | Structural | RT-PCR |  | No |
| *J Clin Microbiol* | Lambert-Niclot S, Cuffel A, Le Pape S, et al. ^65^ | Europe | 138 |  | UR | E, single | Structural | RT-PCR |  | No |
| *J Med Virol* | Infantino M, Grossi V, Lari B, et al. ^66^ | Europe | 61 |  | Serum | S, N | Structural | RT-PCR /CLIA | IgM + IgG | Yes |
| *Euro Surveill* | Jääskeläinen AJ, Kekäläinen E, Kallio-Kokko H, et al. ^67^ | Europe | 37 |  | Serum | N, E, RdRp | Both | RT-PCR /ELISA | IgA + IgG | Yes |
| *J Infect* | Tré-Hardy M, Blairon L, Wilmet A, et al. ^68^ | Europe | 182 |  | Serum |  |  | RT-PCR /ELISA + CLIA | IgA + IgG | No |
| *Orvo Hetil* | Vásárhelyi B, Kristóf K, Ostorházi E, Szabó D, Prohászka Z, Merkely B. ^69^ | Europe | 2310 | 0.06 | UR + Serum |  |  | RT-PCR /LFIA | IgM + IgG | Yes |
| *Infect Ecol Epidemiol* | Hoffman T, Nissen K, Krambrich J, et al.^70^ | Europe | 153 |  | Serum |  |  | RT-PCR /LFIA | IgM + IgG | Yes |
| *J Med Virol* | Cassaniti I, Novazzi F, Giardina F, et al. ^71^ | Europe | 110 |  | UR + Serum | E, RdRp | Both | RT-PCR /LFIA | IgM + IgG | No |
| *medRxiv* | Garcia FP, Perez Tanoira R, Romanyk Cabrera JP, et al. ^72^ | Europe | 100 |  | Serum | N, ORF1Ab | Both | RT-PCR /LFIA | IgM + IgG | Yes |
| *medRxiv* | Paradiso AV, De Summa S, Loconsole D, et al.^73^ | Europe | 190 |  | UR + Serum | N, RNAse P | Both | RT-PCR /LFIA | IgM + IgG | Yes |
| *Public Health* | Döhla M, Boesecke C, Schulte B, et al. ^74^ | Europe | 49 |  | Serum | E, single | Structural | RT-PCR /LFIA | IgM + IgG | Yes |
| *J Emerg Infect Dis* | Okba NMA, Muller MA, Li W, et al. ^75^ | Global | 207 |  | Serum |  |  | RT-PCR /ELISA | IgM + IgG | No |
| *J Clin Virol* | Smithgall MC, Scherberkova I, Whittier S, Green DA. ^76^ | North America | 113 |  | UR | E, ORF1Ab | Both | RT-PCR |  | No |
| *J Med Virol* | Pujadas E, Ibeh N, Hernandez MM, et al. ^77^ | North America | 1006 |  | UR | E, ORF1Ab | Both | RT-PCR |  | No |
| *J Infect Dis* | Burbelo PD, Riedo FX, Morishima C, et al. ^78^ | North America | 100 |  | Serum | N, single | Structural | RT-PCR |  | No |
| *Am J Obstet Gynecol MFM* | Penfield CA, Brubaker SG, Limaye MA, et al. ^79^ | North America | 32 |  | UR + Other | N, single | Structural | RT-PCR |  | No |
| *medRxiv* | Wyllie AL, Fournier J, et al. ^80^ | North America | 44 |  | UR + Other | N, single | Structural | RT-PCR |  | No |
| *J Appl Lab Med* | Suhandynata RT, Hoffman MA, Kelner MJ, McLawhon RW, Reed SL, Fitzgerald RL. ^81^ | North America | 235 |  | Serum | N, single | Structural | RT-PCR /CLIA | IgM + IgG | No |
| *Clin Chem* | Tang MS, Hock KG, Logsdon NM, et al. ^82^ | North America | 201 |  | UR + LR + Serum | N, E | Both | RT-PCR /CLIA + ELISA | IgG | Yes |
| *medRxiv* | Randad PR, Pisanic N, Kruczynski K, et al. ^83^ | North America | 493 |  | UR + Serum + Other | N, single | Structural | RT-PCR /ELISA | IgM + IgA + IgG | No |
| *JMIR Public Health Surveill* | Sullivan PS, Sailey C, Guest JL, et al. ^84^ | North America | 159 |  | UR + Serum | S, N | Structural | RT-PCR /ELISA | IgM + IgA + IgG | No |
| *bioRxiv* | Freeman B, Lester S, Mills L, et al.^85^ | North America | 618 |  | UR NP/OP + Serum | N, single | Structural | RT-PCR /ELISA | IgM + IgG | Yes |

**Fig S1. PRISMA Flowchart for Meta-Analysis and Qualitative Synthesis**

Additional records identified through other sources BioRiXv (871), MedRiXv (3810 ), and Google Scholar(28,050 )
Total (n =32731 )

Studies included in quantitative synthesis (meta-analysis)
(n = 30)

Studies included in qualitative synthesis
(n = 85)

Full-text articles assessed for eligibility
(n = 112)

Records excluded
(n = 24597)

Records screened
(n = 24709)

Records after duplicates removed
(n =24709 )

### Identification

### Eligibility

### Included

### Screening

Full-text articles excluded, duplicated pre-publications
(n = 6), sample size <5 (n =11)

Records identified through PubMed database searching
(n = 2154)

**Suppl. Figure S2.** **Summary plot of risk of bias for each study included in meta-analysis according to QUADAS-2 domain.**

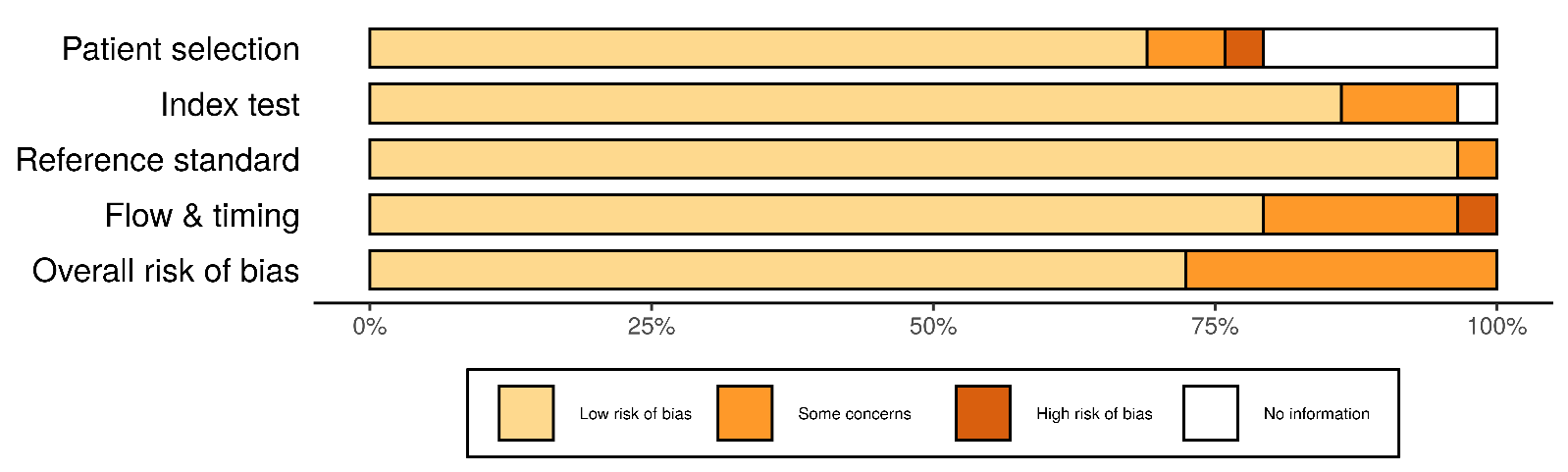
